# Impact of OSA Treatment Success on Changes in Hypertension and Obesity: A Retrospective Cohort Study

**DOI:** 10.1101/2022.04.18.22273955

**Authors:** Akash Shanmugam, Zachary O. Binney, Courtney B. Voyles, Emerson Bouldin, Nikhila P. Raol

**Affiliations:** Emory College of Arts & Sciences, Atlanta, GA, USA; Oxford College of Emory University, Oxford, GA, USA; Children’s Healthcare of Atlanta, Atlanta, GA, USA; Department of Otolaryngology-Head and Neck Surgery, Emory University School of Medicine, Atlanta, GA, USA

**Keywords:** obstructive sleep apnea, hypertension, obesity, treatment outcomes, children

## Abstract

**Objective:** Pediatric obstructive sleep apnea (OSA) has been shown to lead to the development of chronic cardiometabolic conditions, including obesity and cardiovascular disease. We sought to describe the impact of the success of continuous positive airway pressure (CPAP) and surgery, common treatment options for pediatric OSA, on cardiometabolic conditions.

**Methods:** A retrospective review of patients (≤18 years) diagnosed with OSA based on a polysomnogram at a tertiary care pediatric otolaryngology practice from 2015 to 2019 was conducted. Clinical data, including the systolic blood pressure (SBP) values, body mass index (BMI), overall apnea/hypopnea index (AHI) values, and CPAP compliance, were collected. Linear mixed-effects models were developed to observe the relationship between the clinical measurements of each comorbidity and OSA treatment modalities.

**Results:** 507 patients were included. BMI and SBP measures were collected for 230 and 277 patients respectively. The difference-in-difference estimate for the SBP z-score percentile after successful treatment was -5.3 ± 2.0 percentile units per 100 days. The difference-in-difference estimate for SBP z-score percentile after successful CPAP treatment was -14.4 ± 4.9 percentile units per 100 days while the estimate after successful surgical treatment was -4.6 ± 2.3 percentile units per 100 days. No significant differences were found between clinical measures for obese patients in any treatment cohort.

**Conclusions:** The success of OSA management was shown to have a positive impact on SBP in hypertensive patients and no impact on BMI in obese patients. In hypertensive patients, CPAP success tripled improvements in SBP z-score percentile compared to surgical treatment success.

## 1 Introduction

Pediatric obstructive sleep apnea (OSA) is a physiologically disruptive disorder that is characterized by increased pharyngeal airway resistance that ranges from habitual snoring to complete airway obstruction during sleep.^1^ Pediatric OSA is estimated to affect approximately 1% of the entire pediatric population, yet 90% of those children who suffer from sleep disorders are often undiagnosed for a significant period of time. ^2,3^ From the first year after OSA diagnosis, children with OSA experience a 215% increase in healthcare utilization due to OSA-associated morbidity.^4^ The burden of pediatric OSA has also been associated with poor school performance and higher risks for life-long cognitive impairment.^5,6^

Pediatric OSA frequently accompanies or leads to chronic cardiometabolic conditions, including obesity and cardiovascular disease.^7,8^ Effective management of pediatric OSA is critical because hypoxia and poor sleep quality are believed to play crucial roles in the pathogenesis of OSA-related comorbidities.^9^ The impact of pediatric OSA treatment success on cardiometabolic condition of patients with comorbidities has not been well described. While adenotonsillectomy is the first-line treatment for the improvement of sleep-disordered breathing, CPAP has been shown to be more effective at the resolution of residual OSA, especially in patients with metabolic dysfunction.^10,11^A determination of effectiveness of OSA treatments for patients with specific comorbidities could be critical to the development of guidelines that improve pediatric outcomes from chronic illnesses.^12^

While individual treatments have been shown to impact cardiometabolic conditions in patients, there are no studies that compare the changes in clinical measures of different treatments on the comorbidities of OSA. This retrospective study aims to answer the following questions: 1) How does successful pediatric OSA treatment impact hypertension and obesity in comorbid patients? 2) Is there a difference in the changes in hypertension and obesity between surgery and CPAP as treatment options for the management of OSA? In order to answer these questions, clinical and demographic data from pediatric patients with OSA at a tertiary care children’s hospital were studied to examine the effect of OSA management on BMI and systolic blood pressure z-score percentile over time.

## 2 Methods

### 2.1 Inclusion Criteria

Children’s Healthcare of Atlanta Institutional Review Board approval was obtained. A retrospective cohort study of all pediatric patients (≤18 years) who received a polysomnogram at a tertiary care pediatric otolaryngology practice from January 3, 2015 to December 31, 2019 was conducted. Demographic information, including sex, age, and race, was collected. Clinical information about comorbidities, including the systolic blood pressure values, height, weight, and all their respective dates of measurement, were collected over the study’s observation period. Patients who did not have a clinical measurement before and after the start of their treatment were excluded. Polysomnogram data was collected once within six months prior to treatment and again within one year after treatment, and compliance data was collected after usage of CPAP.

Patients who did not have a polysomnogram before and after surgery, a polysomnogram before the start of CPAP use, or information on CPAP compliance, were excluded.

### 2.2 Comorbidity and Treatment Success Classifications

The presence of comorbidities in each patient was categorized by the abnormal levels for each clinical measurement of the disease. The height and weight measurements were converted to body mass index (BMI) and BMI z-score percentile based on the Center for Disease Control (CDC) growth charts^13^. Obese patients were categorized as patients with a BMI z-score percentile measurement above the 95^th^ percentile. Systolic blood pressure (SBP) values were converted to z-score percentile based on the National Institutes of Health (NIH) charts^14^. Patients with hypertension and cardiovascular risk were categorized as patients with a systolic blood pressure z-score measurement above the 95^th^ percentile. Obese patients and hypertensive patients were split into separate cohorts to understand the association of OSA treatment success with BMI and SBP Z-score percentiles respectively.

The success of pediatric OSA treatment was categorized by a substantial decrease in the apnea-hypopnea index (AHI) after surgery or consistent compliance with CPAP treatment. Success for surgical treatment of OSA was classified as either a post-treatment AHI that was less than 5 events per hour or a 50% decrease in AHI if postoperative AHI was >5 events per hour. Success or compliance for CPAP as treatment for OSA was classified as at least 4 hours of CPAP usage a night for 70% of nights in any given time frame. Patients who received treatment for their OSA but did not qualify for these standards of treatment success were considered to have failed treatment.

### 2.3 Statistical Analysis

Linear mixed-effects models were developed to observe the relationship between the success of any OSA treatment and clinical measurements of the cardiometabolic condition of patients with comorbidities. Linear mixed-effects models were also developed to observe the relationship between the success of either CPAP or surgery as treatment for patients’ OSA and clinical measurements of the cardiometabolic condition of patients with the comorbidities. The linear mixed-effects model included a random effect that accounted for differences between patients and fixed effects that controlled for sex, age, race, and pre-treatment severity of sleep apnea. The linear mixed model for hypertensive patients also included a fixed effect for the BMI z-score percentile. The β-coefficients were used to estimate the linear slopes of clinical values before and after OSA treatment and their intercepts. All analyses were performed in R Version 4.0.0^40^ using tidyverse^41^ and lme4^42^.

### 2.4 Clinical Significance

The average percent change in weight and the absolute change in SBP after treatment from baseline was also calculated by comparing the most recent measurement prior to treatment and the lowest measurement after treatment. Both measures helped to evaluate the clinical significance of OSA treatment success on the respective comorbidities of hypertensive and obese patients.

## 3 Results

### 3.1 Demographics

Over the four-year study period, 230 pediatric patients who were above the 95^th^ percentile for BMI and had a polysomnogram with an overall apnea-hypopnea index (AHI) over 1.0 prior to treatment were identified. BMI measures were collected at least once before and after the polysomnogram. The median period of observation, or the days between the first and last recorded comorbidity measure for each patient, was 240 days (IQR 147, 322). Among the obese patients, 66.5% received successful surgical or CPAP treatment for sleep apnea, while 33.5% of obese patients were considered to have failed treatment. With the exception of age of the patients, all differences between the two groups including observation period, sex, race, and various polysomnography measures, were not significant (p>0.05). See Table 1 for additional demographic details.

**Table 1.** Demographics of Obese Patients with Diagnosed Sleep Apnea

|  | Total Obese<br>Patients<br>(N = 230) | Failed<br>Treatment<br>(N = 77) | Successful<br>Treatment<br>(N = 153) | P-value |
| --- | --- | --- | --- | --- |
| **Treatment Cohorts** |  |  |  |  |
| CPAP | 73 (31.7%) | 49 (67.1%) | 24 (32.9%) |  |
| Surgery | 157 (68.3%) | 28 (17.8%) | 129 (82.2%) |  |
| **Age** | | | | $P = 0.00062^*$ |
| median (IQR) | 9.7<br>(5.0, 14.4) | 12.2<br>(7.2, 16.0) | 8.5<br>(4.4, 13.5) |  |
| **Pre-Treatment BMI**<br>**Z-Score Percentile** | | | | $P = 0.22$ |
| median (IQR) | 99.0<br>(96.6, 99.6) | 99.3<br>(97.6, 99.6) | 98.9<br>(96.3, 99.6) |  |
| **Observation Period in Days** | | | | $P = 0.71$ |
| median (IQR) | 240.0<br>(147, 322) | 242<br>(156, 325) | 240<br>(147, 322) |  |
| **Polysomnography Measures**<br>**(median, IQR)** |  |  |  |  |
| Pre-Treatment Overall AHI | 13.2<br>(6.5, 27.1) | 12.2<br>(6.7, 24.7) | 13.7<br>(6.2, 27.6) | $P = 0.75$ |
| Lowest Oxygen Saturation | 83.0<br>(76.1, 88.0) | 83.0<br>(76.5, 88.0) | 84.4<br>(76.0, 88.0) | $P = 0.39$ |
| Percent of Sleep Time in REM | 19.2 | 19.0 | 19.6 | $P = 0.60$ |
|  | (14.0, 24.6) | (11.2, 25.5) | (15.1, 23.8) |  |
| Percent of Sleep Time with<br>end-tidal CO2 values > 50mm Hg | 0.0<br>(0.0, 5.2) | 0.0<br>(0.0, 3.7) | 0.0<br>(0.0, 5.3) | *P* = 0.65 |
| **Sex** |  |  |  | *P* = 0.6739 |
| Male | 133 (57.8%) | 43 (55.8%) | 90 (58.8%) |  |
| Female | 97 (42.2%) | 34 (44.2%) | 63 (41.2%) |  |
| **Race** |  |  |  | *P* = 0.5733 |
| Non-Hispanic White | 59 (25.7%) | 22 (28.6%) | 37 (24.2%) |  |
| Non-Hispanic Black | 110 (47.8%) | 36 (46.8%) | 74 (48.4%) |  |
| Asian | 4 (1.7%) | 0 (0.0%) | 4 (2.6%) |  |
| Hispanic | 46 (20.0%) | 14 (18.2%) | 32 (20.9%) |  |

In the second pediatric cohort, 277 patients were identified who were above the 95^th^ percentile for systolic blood pressure and had a polysomnogram with an overall AHI over 1.0 prior to treatment. The median period of observation was 123 days (IQR 9, 293). Among the hypertensive patients, 75.1% received successful surgical or CPAP treatment for sleep apnea, while treatment did not succeed for 24.9% of obese patients. With the exception of age and observation period of the patients, all differences between the two groups including sex, race, and various polysomnography measures, were not significant (p>0.05). See Table 2 for additional demographic details.

**Table 2.** Demographics of Hypertensive Patients with Diagnosed Sleep Apnea

|  | Total<br>Hypertensive<br>Patients<br>(N = 277) | Failed<br>Treatment<br>(N = 69) | Successful<br>Treatment<br>(N = 208) | P-value |
| --- | --- | --- | --- | --- |
| **Treatment Cohorts** |  |  |  |  |
| CPAP | 38 (13.7%) | 25 (65.8%) | 13 (34.2%) |  |
| Surgery | 239 (86.3%) | 44 (18.4%) | 195 (81.6%) |  |
| **Age** |  |  |  | *\*P* = 0.0045 |
| median (IQR) | 6<br>(3.0, 11.0) | 8<br>(4.0, 13.0) | 5.0<br>(3.0, 9.0) |  |
| **Pre-Treatment BMI**<br>**Z-Score Percentile** |  |  |  | *P* = 0.42 |
| median (IQR) | 89.5 | 93. | 88.8 |  |
|  | (51.4, 98.9) | (69.6, 99.2) | (48.3, 98.7) |  |
| **Observation Period in Days** |  |  |  | *\*P* = 0.0004 |
| median (IQR) | 123<br>(9, 293) | 222<br>(97, 336) | 83.5<br>(8, 253) |  |
| **Polysomnography Measures**<br>**(median, IQR)** |  |  |  |  |
| Pre-Treatment Overall AHI | 13.1<br>(6.4, 24.4) | 11.0<br>(6.8, 19.6) | 13.7<br>(6.2, 24.9) | *P* = 0.29 |
| Lowest Oxygen Saturation | 82.0<br>(74.0, 88.0) | 81.0<br>(74.0, 86.0) | 83.0<br>(74.0, 88.0) | *P* = 0.26 |
| Percent of Sleep Time in REM | 268; 19.0 (14.0, 25.0) | 66; 17.0<br>(11.1, 23.9) | 202; 19.7<br>(14.9, 25.0) | *P* = 0.081 |
| Percent of Sleep Time with<br>end-tidal CO2 values > 50mm<br>Hg | 0.1<br>(0.0, 7.6) | 0.1<br>(0.0, 8.6) | 0.0<br>(0.0, 7.3) | *P* = 0.62 |
| **Sex** |  |  |  | *P* = 0.4008 |
| Male | 161 (58.1%) | 37 (53.6%) | 124 (59.6%) |  |
| Female | 116 (41.9%) | 32 (46.4%) | 84 (40.4%) |  |
| **Race** |  |  |  | *P* = 0.9291 |
| Non-Hispanic White | 87 (31.4%) | 23 (33.3%) | 64 (30.8%) |  |
| Non-Hispanic Black | 126 (45.5%) | 29 (42.0%) | 97 (46.6%) |  |
| Asian | 6 (2.2%) | 2 (2.9%) | 4 (1.9%) |  |
| Hispanic | 49 (17.7%) | 13 (18.8%) | 36 (17.3%) |  |
| **Earliest Pre-Treatment**<br>**Systolic Blood Pressure**<br>**Z-Score Percentile** |  |  |  | *P* = 0.77 |
| median (IQR) | 91.4<br>(60.6, 99.1) | 90.9<br>(57.9, 99.4) | 91.4<br>(67.0, 99.0) |  |

### 3.2 Obese Patients

In order to observe the impact of OSA treatment success on obesity, the BMI z-score percentiles for patients were collected before and after treatment. The linear mixed effects model indicated that success of OSA treatment was not associated with a significant change in BMI z-score percentiles values of patients. The difference-in-difference estimate for the association between OSA treatment success and BMI Z-Score Percentile was 1.2 ± 4.3 percentile points per 100 days. See Figure 1 for the visualization of the linear mixed effects model and Table 3 for additional details on the difference-in-difference estimate.

**Table 3.** Change of BMI Z-Score Percentile After OSA Treatment

| | Change in BMI Z-Score Percentile Before Treatment (percentile points per 100 days, mean ± SE) | | Change in BMI Z-Score Percentile After Treatment (percentile points per 100 days, mean ± SE) | | Difference-in-Difference Estimate Between Cohorts (percentile points per 100 days, mean ± SE) |
| --- | --- | --- | --- | --- | --- |
|  | Treatment Failure | Treatment Success | Treatment Failure | Treatment Success |  |
| All Patients | -0.3 ± 2.6 | 0.1 ± 2.0 | -2.7 ± 1.5 | -1.0 ± 1.0 | 1.2 ± 4.3 |
| CPAP | -1.1 ± 2.8 | 0.2 ± 4.0 | 0.1 ± 1.7 | 0.3 ± 1.9 | -1.1 ± 6.5 |
| Surgery | 0.6 ± 4.1 | -0.01 ± 2.37 | -6.2 ± 2.4 | -1.3 ± 1.2 | 5.5 ± 6.2 |

**Figure 1.**
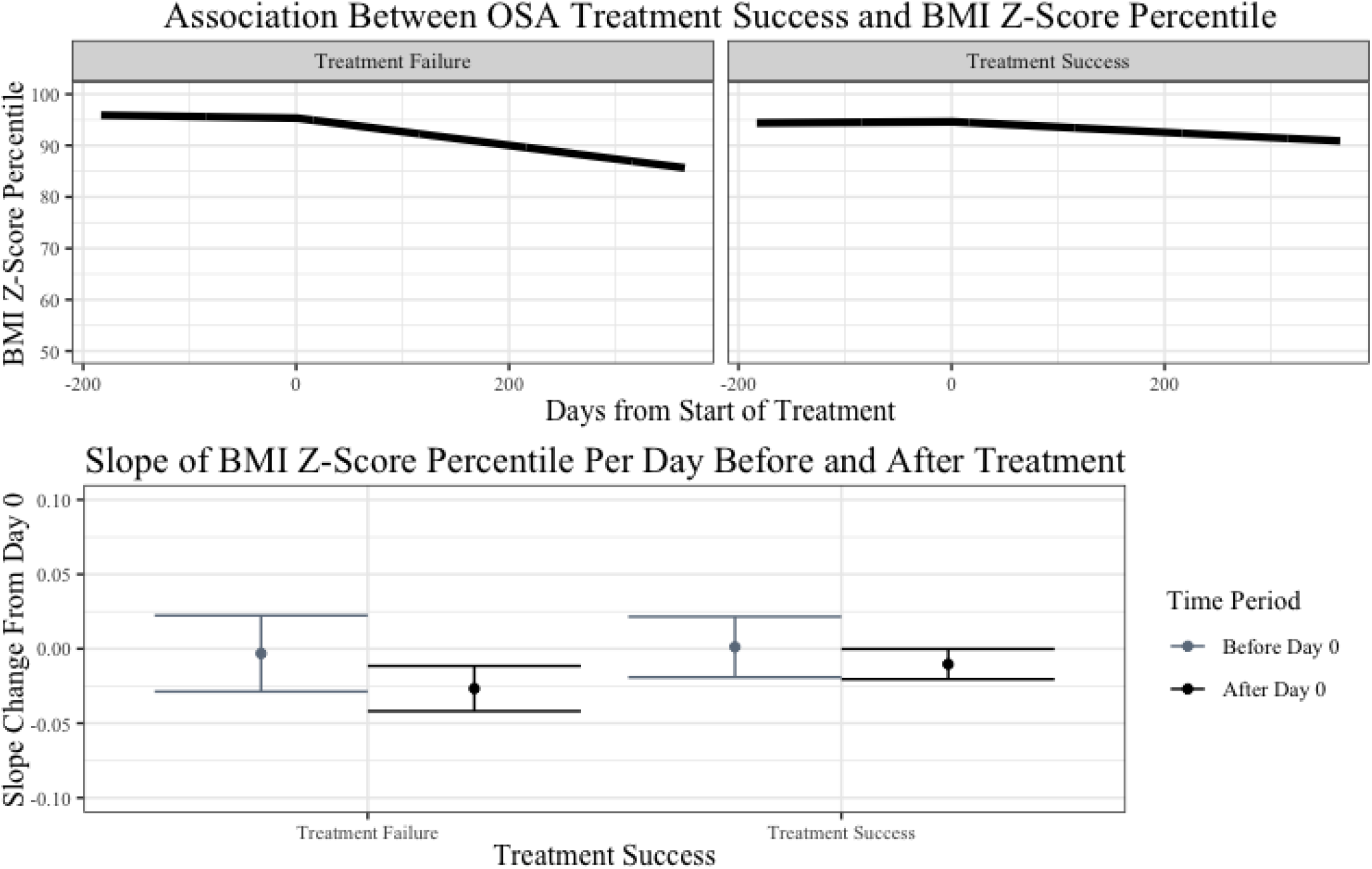
A linear mixed effects model was plotted to estimate the association between pediatric OSA treatment success and BMI Z-Score percentile from 0.5 years prior to treatment to 1 year after treatment (top). The model controlled for the type of treatment, sex, race, age, and the AHI prior to treatment of the patient. The pre-slope and post-slope of the model are shown with confidence intervals (bottom).

After separating patients into CPAP and surgical treatment cohorts, the change of BMI Z-Score percentile over time was also observed. The linear mixed effects model indicated that success of CPAP as treatment for pediatric OSA was not associated with a significant change in BMI Z-Score percentile of patients. The difference-in-difference estimate for the association between OSA treatment success and BMI Z-Score Percentile was -1.1 ± 6.5 percentile points per 100 days. See Figure 2 for the visualization of the linear mixed effects model.

**Figure 2.**
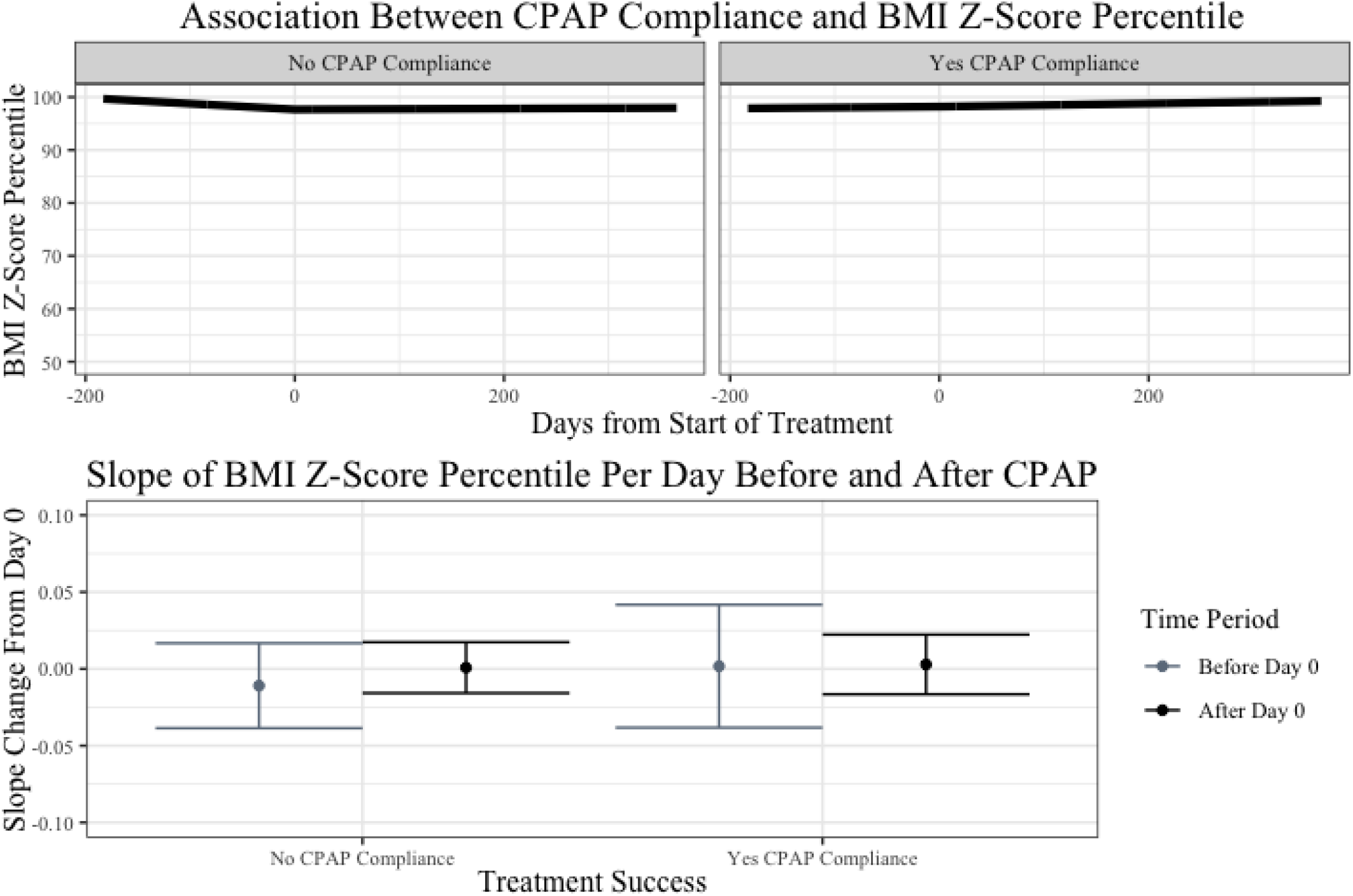
A linear mixed effects model was plotted to estimate the association between pediatric CPAP treatment success and BMI Z-Score percentile from 0.5 years prior to treatment to 1 year after treatment (top). The model controlled for the type of treatment, sex, race, age, and the AHI prior to treatment of the patient. The pre-slope and post-slope of the model are shown with confidence intervals (bottom).

In the surgical cohort for BMI Z-Score Percentile, the linear mixed effects model indicated a significant decrease in the BMI Z-Score percentile for patients who did not have successful surgical treatment for OSA. While BMI Z-Score percentile changed by 0.6 ± 4.1 percentile points per 100 days prior to unsuccessful surgical treatment, BMI Z-Score percentile decreased by 6.2 ± 2.4 percentile points per 100 days after unsuccessful treatment. See Figure 3 for the visualization of the linear mixed effects model and Table 3 for additional details on slopes before and after OSA treatment.

**Figure 3.**
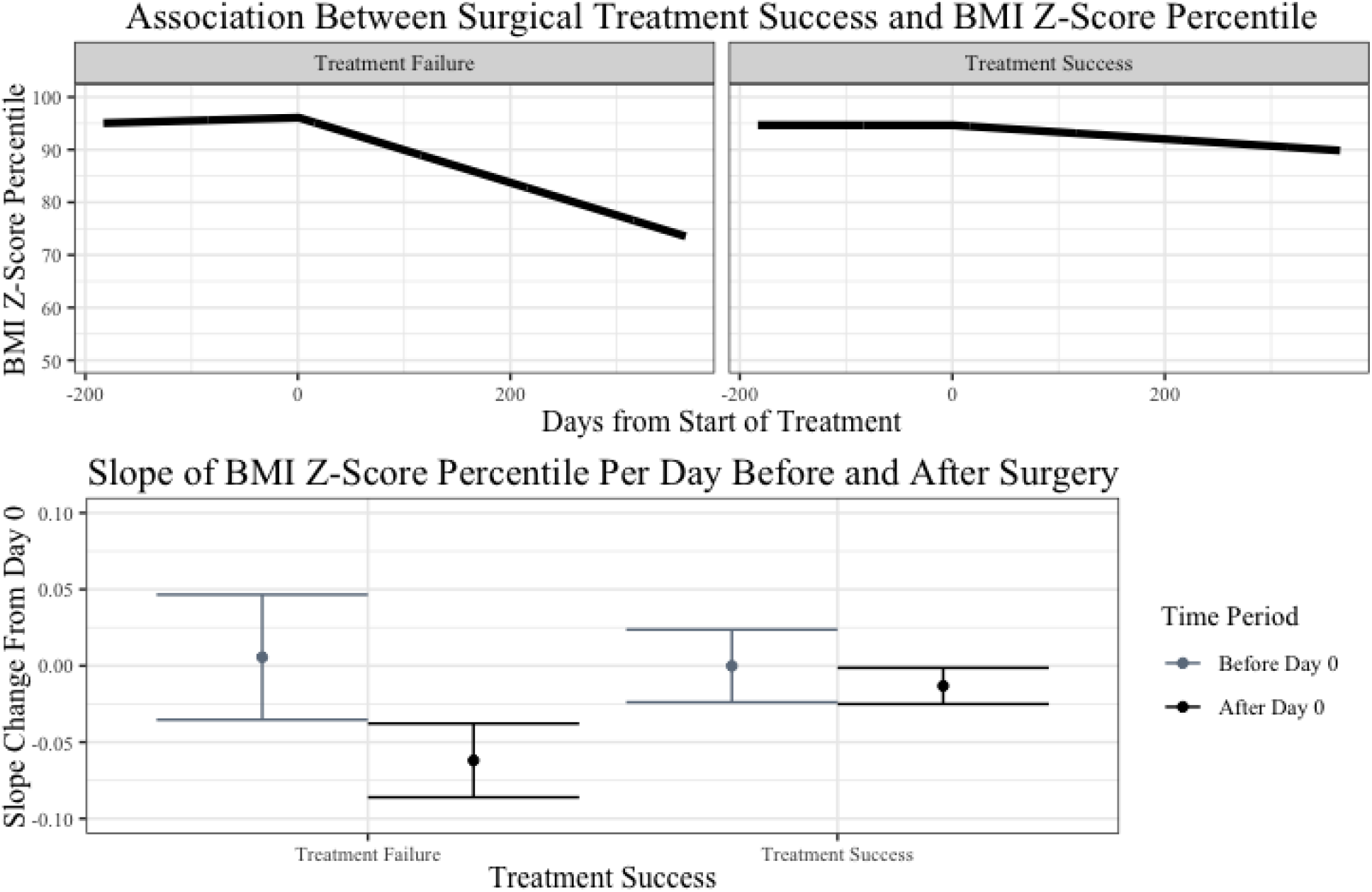
A linear mixed effects model was plotted to estimate the association between pediatric surgical treatment success and BMI Z-Score percentile from 0.5 years prior to treatment to 1 year after treatment (top). The model controlled for the type of treatment, sex, race, age, and the AHI prior to treatment, and the AHI change after treatment. The pre-slope and post-slope of the model are shown with confidence intervals (bottom).

The average percent change in weight was also compared in patients between different treatment modalities. No significant differences were found between successful and unsuccessful treatment among different treatment modalities. See Table 4 for the average percent weight change of patients across treatment modalities.

**Table 4.** Average Percent Change in Weight after OSA Treatment

|  | Failed Treatment | Successful Treatment |
| --- | --- | --- |
| OSA Treatment | $3.5 \pm 3.2\%$ | $1.8 \pm 2.1\%$ |
| CPAP | $7.0 \pm 4.4\%$ | $1.5 \pm 3.2\%$ |
| Surgery | $-0.6 \pm 4.1\%$ | $1.8 \pm 2.4\%$ |

### 3.3 Hypertensive Patients

The linear mixed effects model indicated that OSA treatment was associated with a significant increase in SBP z-score percentile in patients who did not have successful treatment and with a significant decrease in SBP z-score percentile in patients who had successful treatment. Prior to treatment, systolic blood pressure z-score percentiles in both failed and successful treatment cohorts were slightly declining at -1.1 ± 1.2 percentile points per 100 days or staying constant at -0.4 ± 1.0 percentile points per 100 days. After treatment, differences in the slopes of systolic blood pressure z-score percentiles in the failed and successful treatment cohorts emerged. In the failed treatment cohort, systolic blood pressure z-score percentile was associated with a significant increase of 2.7 ± 0.6 percentile points per 100 days while the successful treatment cohort was associated with a significant decrease in systolic blood pressure z-score percentile by -1.9 ± 0.5 percentile points per 100 days. The difference-in-difference estimate for the association between OSA treatment success and SBP z-score percentile was -5.3 ± 2.0 percentile points per 100 days, suggesting that success in OSA treatment was associated with a decrease in SBP z-score percentile. See Figure 4 for the visualization of the linear mixed effects model and Table 5 for the slope values of both treatment success cohorts before and after the start of treatment.

**Table 5.** Change of SBP Z-Score Percentile After OSA Treatment

| | Change in SBP Z-Score Percentile Before Treatment<br>(percentile points per 100 days, mean ± SE) | | Change in SBP Z-Score Percentile After Treatment<br>(percentile points per 100 days, mean ± SE) | | Difference-in-Difference Estimate Between Cohorts<br>(percentile points per 100 days, mean ± SE) |
| --- | --- | --- | --- | --- | --- |
|  | Treatment Failure | Treatment Success | Treatment Failure | Treatment Success |  |
| All Patients | -1.1 ± 1.2 | -0.4 ± 1.0 | 2.7 ± 0.6 | -1.9 ± 0.5 | -5.3 ± 2.0 |
| CPAP | 0.4 ± 2.2 | 5.7 ± 2.4 | 4.0 ± 1.2 | -5.1 ± 2.1 | -14.4 ± 4.9 |
| Surgery | -2.5 ± 1.5 | -1.7 ± 1.1 | 2.2 ± 0.7 | -1.7 ± 0.5 | -4.6 ± 2.3 |

**Figure 4.**
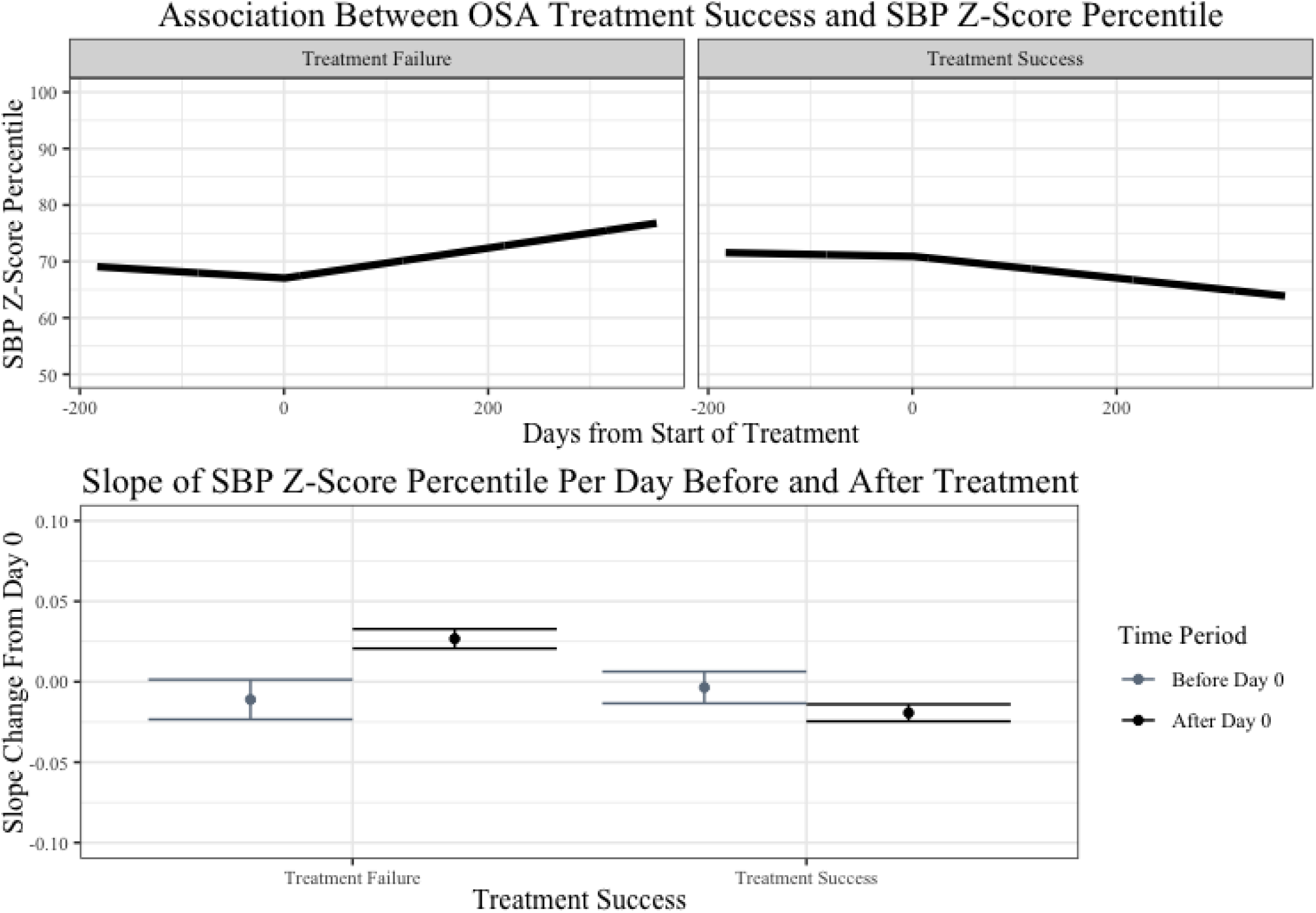
A linear mixed effects model was plotted to estimate the association between the success of pediatric OSA treatment and systolic blood pressure z-score percentile from 0.5 years prior to treatment to 1 year after treatment (top). The model controlled for the type of treatment, sex, race, age, BMI Z-Score of the patient, and the AHI prior to treatment of the patient. The pre-slope and post-slope of the model are shown with confidence intervals (bottom).

In the cohort of patients who used CPAP as treatment, the linear mixed-effects model indicated that CPAP treatment was associated with a significant increase in systolic blood pressure z-score percentile for patients who were not compliant with treatment and a significant decrease for patients who were compliant with treatment. Prior to treatment, patients who were not compliant with CPAP treatment saw no change in their systolic blood pressure z-score percentile at 0.4 ± 2.2 percentile points per 100 days. On the other hand, patients who were compliant with CPAP treatment experienced an increase in their systolic blood pressure z-score percentile prior to treatment at 5.7 ± 2.4 percentile points per 100 days. However, both groups experienced significant changes in their systolic blood pressure z-score percentile after the start of treatment. Patients who were not compliant with CPAP treatment experienced an increase of 4.0 ± 1.2 percentile points per 100 days while patients who were compliant with CPAP treatment experienced a decrease of 5.1 ± 2.1 systolic blood pressure z-score percentile points per 100 days. The difference-in-difference estimate for success of CPAP treatment was -14.4 ± 4.9 percentile points per 100 days, indicating that the success of CPAP treatment was associated with a substantial decrease in SBP z-score percentile. See Figure 5 for the visualization of the linear mixed effects model and Table 5 for the slope values of both treatment success cohorts before and after the start of treatment.

**Figure 5.**
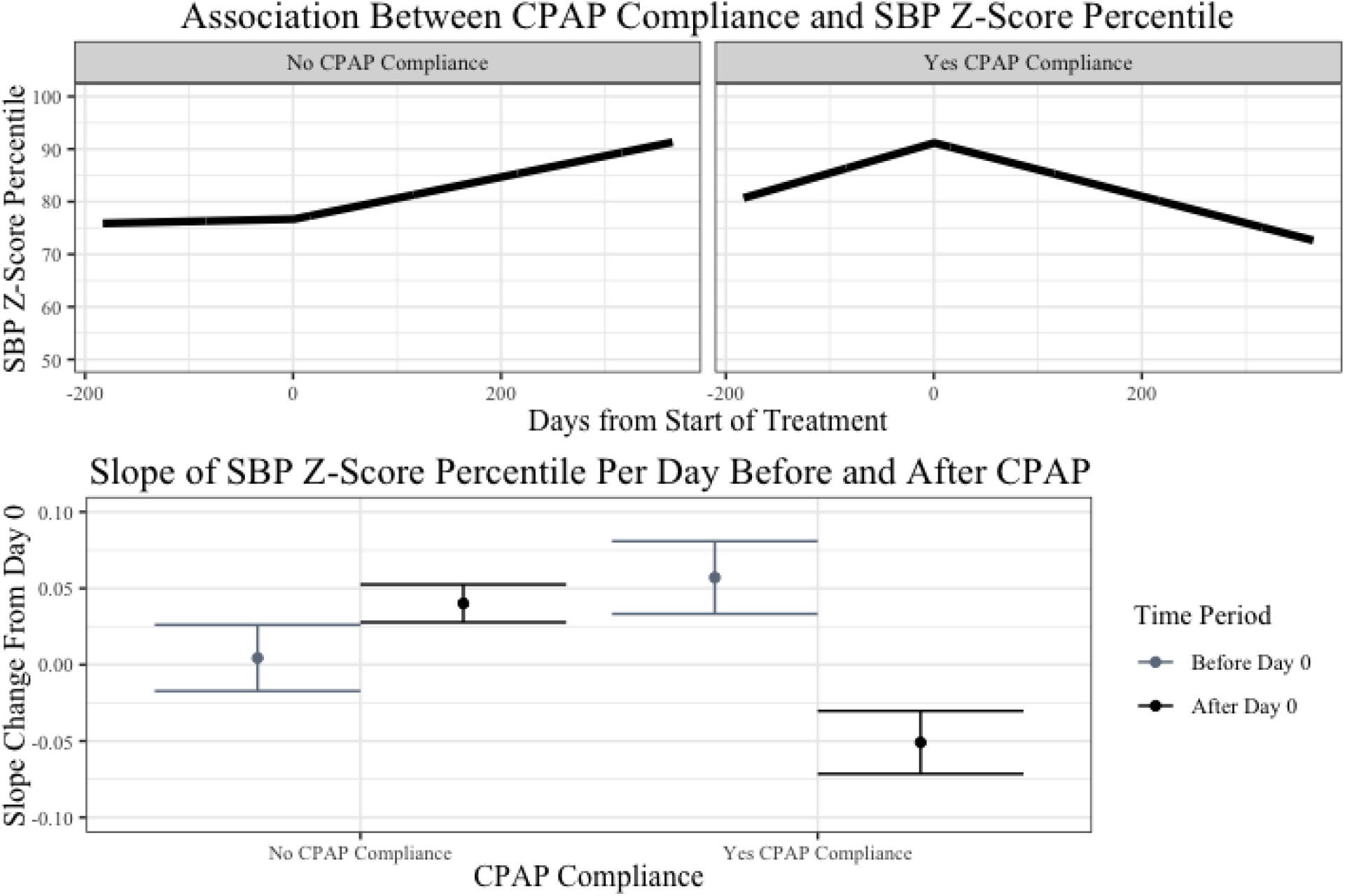
A linear mixed effects model was plotted to estimate the association between CPAP compliance and systolic blood pressure z-score percentile from 0.5 years prior to treatment to 1 year after treatment (top). The model controlled for the type of treatment, sex, race, age, BMI Z-Score of the patient, and the AHI prior to treatment of the patient. The pre-slope and post-slope of the model are shown with confidence intervals (bottom).

In the patient cohort who used surgical treatment, the linear mixed-effects model indicated an increase in systolic blood pressure z-score percentile after treatment in patients who failed their treatment program but no change in systolic blood pressure z-score percentile after treatment in patients who had successful surgical treatment for their OSA. Prior to treatment, the systolic blood pressure z-score percentile of both groups of patients who failed and successful treatment was decreasing at -2.5 ± 1.5 and -1.7 ± 1.1 percentile points per 100 days. After surgical treatment, patients whose surgical treatment failed to significantly improve their AHI were associated with a significant increase in systolic blood pressure z-score percentile by 2.2 ± 0.7 percentile points per 100 days. Patients whose surgical treatment improved their AHI were associated with a continued decrease of systolic blood pressure z-score percentile by -1.7 ± 0.5 percentile points per 100 days. For patients with successful treatment, the post-treatment slope was not significantly different from the pre-treatment slope. The difference-in-difference estimate for success of surgical treatment was -4.6 ± 2.3 percentile points per 100 days, indicating that the success of surgery was associated with a modest decrease in SBP z-score percentile. See Figure 6 for the visualization of the linear mixed effects model and Table 5 for the slope values of both treatment success cohorts before and after the start of treatment.

**Figure 6.**
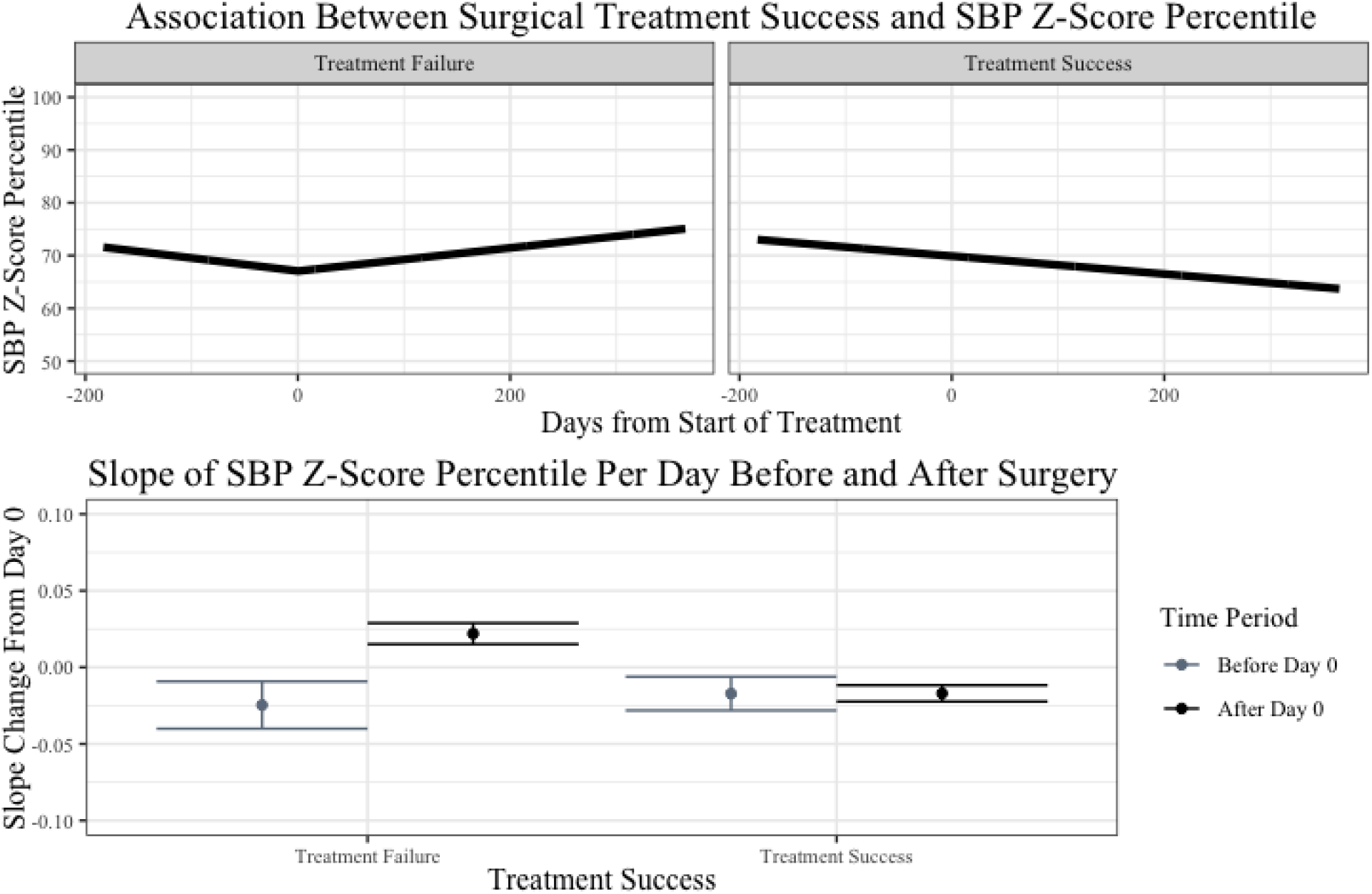
A linear mixed effects model was plotted to estimate the association between surgical treatment success and systolic blood pressure z-score percentile from 0.5 years prior to treatment to 1 year after treatment (top). The model controlled for the type of treatment, sex, race, age, BMI Z-Score of the patient, the AHI prior to treatment, and the AHI change after treatment. The pre-slope and post-slope of the model are shown with confidence intervals (bottom).

The average percent change in systolic blood pressure was also compared between different treatment modalities. No significant differences were found between successful and unsuccessful treatment. See Table 6 for the average percent weight change of patients across treatment modalities.

**Table 6.** Average Change in Systolic Blood Pressure (mmHg) for Each Cohort

|  | Failed Treatment | Successful Treatment |
| --- | --- | --- |
| OSA Treatment | $1.6 \pm 3.3$ | $-2.4 \pm 2.6$ |
| CPAP | $4.0 \pm 3.7$ | $-2.5 \pm 10.4$ |
| Surgery | $0.5 \pm 4.7$ | $-2.4 \pm 2.7$ |

## 4 Discussion

Pediatric obstructive sleep apnea (OSA), a physiologically disruptive disorder, has been believed to play a crucial role in the pathogenesis of chronic cardiometabolic conditions, like obesity and cardiovascular disease.^7,8,9^ This retrospective study aims to answer the following questions: What is the impact of pediatric OSA treatment success on clinical measures of hypertension and obesity in comorbid patients? Is there a difference between surgery and CPAP as treatment options for the management of hypertension and obesity in these comorbid patients?

### 4.1 Obese Patients

The BMI Z-score percentile of obese patients were compared over time between those who experienced success of OSA treatment and those who were considered to have failed OSA treatment. This effect was also compared between pediatric patients who used surgery and those who used CPAP as treatment for their OSA. The success of any OSA treatment was not associated with a significant change in BMI z-score percentile of obese patients. The success of either surgery or CPAP as treatment for OSA also were not associated with any significant change in BMI z-score percentile of obese pediatric patients. Our results also indicated that the differences of the average percent change in weight between patients who succeeded and failed treatment were not significant for any cohort.

The non-significant impact of surgical success for pediatric OSA is consistent with the findings of previous retrospective and prospective studies on weight changes in obese children after an adenotonsillectomy, a common surgical option for OSA. Most retrospective studies did not find any significant effect of an adenotonsillectomy on weight gain^13^, BMI^16^, BMI z-score^17^, or BMI range^18^ in obese children. One retrospective study that did find a significant effect of adenotonsillectomy on BMI in obese children was limited by a small sample size and only included 7 obese children.^19^ One study performed secondary analysis on prospective data from Childhood Adenotonsillectomy Trial (CHAT), a multi-center, randomized controlled trial, and did not find a significant association between an adenotonsillectomy and an undesirable increase in BMI z-score in obese children.^20^ However, limited research has focused on the impact of CPAP use on clinical measures of pediatric obesity.^21^ This is the first study that compares the success of CPAP use on BMI z-score percentile in the pediatric population.

One surprising result was that the patients who were considered to have failed surgery as treatment for OSA were associated with a significant decrease in change in BMI Z-score percentile after treatment. Several studies have recommended bariatric surgery as a first-line surgery or weight-loss intervention programs to treat pediatric OSA in obese pediatric patients^22,23,24^. The weight loss after failed surgical treatment during this study’s observation period could be explained by physicians utilizing an additional intervention aimed at weight loss to resolve sleep-disordered breathing in the patients.

While OSA and obesity can individually negatively impact on quality of life, both illnesses can coexist and potentiate their damaging effects on pediatric health.^7^ The concurrent presence of both obesity and OSA in the pediatric population has been shown to markedly increase the risk of endothelial dysfunction and cardiovascular risk factors compared to the presence of either illness individually.^25^ Because of the intimate relationship of obesity and OSA in the pediatric population, this study’s consideration of the impact of OSA treatment success on BMI z-score percentile change after treatment has value.

Our results indicate that the success of any OSA treatment, CPAP, or surgery, does not have a significant association with a decrease in BMI z-score percentile in obese patients. This suggests that BMI is a mediator for OSA, rather than the inverse relationship between OSA treatment success and BMI change. This is supported by other findings on the relationship of BMI and change in AHI. A population-based study indicated that obesity was the most significant risk factor for pediatric OSA and suggested a 12% increase in risk for OSA for each unit increase in BMI.^26^ A retrospective chart review also indicated an inverse linear relationship between BMI z-score and improvement in total AHI after adenotonsillectomy compared to a control group.^27^ Because OSA treatment success on its own does not have a significant impact on BMI z-score percentile, OSA treatment should be coupled with a weight loss intervention program to resolve both obesity and OSA in pediatric patients with those comorbidities. However, because the findings are limited by the retrospective nature of the study, controlled prospective studies should be conducted on the effectiveness of the suggested intervention before use in the pediatric population.

### 4.2 Hypertensive Patients

For patients with elevated blood pressure classified as hypertension, success of any OSA treatment was associated with a significant decrease in systolic blood pressure z-score percentile change over time after treatment while the failure of any OSA treatment was associated with a significant increase after treatment. The difference-in-difference estimates for overall OSA treatment success and surgical success on systolic blood pressure z-score percentile both indicated modest decreases, while the estimate for CPAP success indicated a more substantial decrease and was three times higher than the estimate for surgical success. The surgical treatment success cohort might be limited by a floor effect because its pre-treatment SBP z-score percentile slope was lower than the pre-treatment slope of CPAP compliance success. Without additional evidence to support this effect, CPAP is likely to be a better treatment option for management of OSA in hypertensive pediatric patients. These differences between cohorts were not shown to be clinically significant as the average change in SBP was not significantly different among different treatment modalities. In the adult population, a 10 mm Hg change has been associated with a 22% reduction in the risk for coronary heart disease and 41% reduction in the risk for a stroke.^28^ However, it is unclear if OSA treatment success has this substantial effect on systolic blood pressure.

These findings align with studies that retrospectively or prospectively measure SBP after surgical treatment on OSA in hypertensive patients in the pediatric population. Three retrospective studies observed SBP in hypertensive OSA pediatric patients 24 hours after adenotonsillectomy using ambulatory blood pressure monitoring and found a significant decrease in SBP after treatment.^29,30,31^ Another retrospective study observed SBP in hypertensive OSA pediatric patients who underwent surgical treatment for OSA after a median observation period of 10 months and found a significant decrease in systolic blood pressure after treatment.^32^ A prospective study compared hypertensive and non-hypertensive pediatric patients both 24 hours and 6 months after adenotonsillectomy and found a significant decrease of ambulatory SBP in hypertensive pediatric patients.^33^ Our findings that surgical treatment success was associated with a continued decline in SBP z-score percentile in hypertensive patients aligned with the results of these studies. The finding on the improvement of SBP z-score percentile after CPAP compliance success aligned with a retrospective study that evaluated CPAP usage on SBP in OSA patients and found a significant decrease in SBP after 6 months.^34^ Beyond this retrospective study, the impact of CPAP compliance on systolic blood pressure in pediatric OSA patients has not been thoroughly examined.

The relationship between pediatric OSA and hypertension have been investigated to understand the impact of the frequent arousals and disturbances in homeostatic gas exchange common to sleep disordered breathing on cardiovascular outcomes later in life.^35^ A population-based cohort study found that persistent sleep apnea is associated with a threefold increase in elevated blood pressure in adolescence.^36^ Because hypertension presents long-term cardiovascular risks to pediatric patients, this study’s understanding of the impact of OSA treatment success on SBP z-score percentile change after treatment is critical to efficacious treatment of hypertensive pediatric OSA patients.

The severity of pediatric OSA has been associated with increased sympathetic activity overnight, causing persistent increases in vascular resistance and altering blood pressure^37^. Both surgical treatment and CPAP usage for pediatric OSA has been shown to reversibly alter systemic levels of inflammatory biomarkers, reducing the cardiovascular risk of OSA^38,39^. A reduction in sympathetic activity and level of inflammatory biomarkers could have led to the reduction in systolic blood pressure z-score percentile seen in patients who experienced successful treatment. Prospective studies are needed to confirm the efficacy of CPAP or surgery in reducing systemic inflammation and managing both OSA symptoms and systolic blood pressure in hypertensive pediatric OSA patients.

### 4.3 Limitations

Our study retrospectively observed cardiometabolic changes in a large number of participants over a long period before and after treatment with no research intervention in their lives. However, this study has a few limitations. This study characterized pediatric patients at a single institution, which may constrain the generalizability of our findings. Given our diverse population and variety of widely used treatments for pediatric OSA in the analysis, we believe our findings should be applicable to pediatric OSA patients in hospitals across the United States. Our study only included patients who elected to receive treatment for OSA and did not observe the few patients who chose to forego treatment in favor of watchful waiting, because of parental choice, or other reasons. Because of this, our study could not have a control group and instead compared clinical measurements of comorbidities between patients who experienced successful treatment to those who experienced unsuccessful treatment. Because our study aimed to compare the impact of successful and non-successful treatment, our lack of a control group consisting of patients who did not receive any treatment does not cause confounding or impact the internal validity.

Given the retrospective nature of our study, the group assignments are prone to selection bias. Although patients were not randomly assigned to a treatment cohort, our study compared patients who succeeded or failed within each type of treatment, so the lack of random assignment was not an issue in our analysis. While our findings should be generalized with caution, the lack of random assignment should not have biased our findings about patients who received CPAP or surgery. Our study was also prone to possible confounding by the race and socioeconomic status of patients. Our study found no significant differences in the race compositions of either cohort among patients who experienced successful or unsuccessful treatment, and race was also controlled against in the models. However, socioeconomic status was not consistently recorded in health data, so socioeconomic status could not be controlled against in the models or analyzed to understand differences in cohorts. Children with developmental abnormalities were also included in the study, but their comorbidities would not affect levels of obesity or hypertension independently to be considered as a confounder. To control for possible baseline differences between OSA patients and other variables that were not considered, our analysis used a random effects model to adjust for individual differences between patients.

Because the data were retrospectively collected from patient medical records, our analysis could also have unintentionally excluded patients whose health information was miscoded and inaccurately recorded. Patients could also have been included in the wrong cohort based on inaccurate polysomnogram data and treatment records. Although errors in collection of health information occur, physicians and scribes are likely to catch these errors during follow-up visits with patients and either correct them or indicate that there are incorrect measures in the charts. The polysomnogram data and findings from patients who used CPAP as treatment for their OSA are also limited. While a majority of the compliance data was directly taken from CPAP machines, some patient compliance data was self-reported or taken from patient charts. The reporting bias of patients for their CPAP use could be a possible source of exposure misclassification in the analysis of the observed impact of CPAP therapy on the cardiometabolic condition. However, this issue appeared in only a small number of patients.

## 5 Conclusions

While the successful management of pediatric OSA was not associated with a significant effect on clinical measures for obesity, successful management of pediatric OSA was associated with a decrease of systolic blood pressure z-score percentile after treatment. Between CPAP and surgery as OSA treatment options, CPAP was shown to be associated with a larger decrease in systolic blood pressure z-score percentile change over time, indicating its potential value as treatment for hypertensive-OSA patients. Future research should look at the reason for the differential improvement in SBP and help identify which patients are better CPAP vs. surgical candidates.

## Data Availability

All data produced in the present study are available upon reasonable request to the authors.

## Abbreviations

OSA: obstructive sleep apnea
AHI: apnea-hypopnea index
BMI: body mass index
SBP: systolic blood pressure
CPAP: continuous positive airway pressure

